# A Fractional Order Dengue Fever Model in the Context of Protected Travellers

**DOI:** 10.1101/2021.01.09.21249522

**Authors:** E. Bonyah, M. L. Juga, C. W. Chukwu, Fatmawati

## Abstract

Climate changes are affecting the control of many vector-borne diseases, particularly in Africa. In this work, a dengue fever model with protected travellers is formulated. Caputo-Fabrizio operator is utilized to obtain some qualitative information about the disease. The basic properties and the reproduction number is studied. The two steady states are determined and the local stability of the states are found to be asymptotically stable. The fixed pointed theory is made use to obtain the existence and uniqueness of solutions of the model. The numerical simulation suggests that the fractional-order affects the dynamics of dengue fever.

## 1 Introduction

Dengue fever is a mosquito-borne infection in the tropical and sub-tropical regions of the globe. it is caused by any of the four serotypes of the Dengue virus namely: DEN 1, DEN 2, DEN 3 and DEN 4. An estimate of about 50 million people gets infected with the disease every year in about 100 countries. [1]. Infected individuals remain asymptomatic for about 3-14 days (an average of 4-7 days) before they begin to experience a sudden onset of fever. There is no specific cure for Dengue fever disease, hospital treatments include sufficient bed rest, antipyretics, and analgesics given for supportive care. It is believed that dengue virus is quickly cleared in the human body by the natural immune system within approximately 7 days after the day of sudden onset of fever [2, 3].

Several mathematical models have been developed to understand the transmission dynamics of Dengue fever disease. Esteva and Vargas formulated an *SIR* model analyse the dengue fever transmission dynamics with a constant human population [4] and a variable human population [5]. In [6], Edy and Supriatna developed a dengue fever transmission model which restricted the dynamics for the constant host and vector populations. Many other studies have been carried out to investigate the transmission of dengue fever, [7–11]. However, all the different modelling approaches used in these models are limited due to the local nature of integer order derivatives. Fractional calculus in recent times has attracted many researchers because of the memory effect in prediction. Mathematical modelling has become an indispensable tool in providing qualitative information on phenomena in the absence of real data. In 2015, Caputo and Fabrizio developed a novel operator namely the Caputo– Fabrizio (CF) fractional operator which involves a non-singular kernel [12]. Losada and Nieto later reported some additional properties of this operator [13]. Since then, several mathematical models have been developed by researchers using the fractional-order approach [14–18]. These models have demonstrated both the efficiency and suitability of the CF operator. We, therefore propose a CF fractional-order system to analyse the dynamics of Dengue fever in the context of protected travellers.

The rest of the paper is organised as follows: In section 2, we give the preliminaries and define the fractional-order model in section three. In Section 4, we look at the model’s basic properties including the existence and uniqueness of solutions, equilibrium points and their stability analyses. We simulate the model in Section 5 and draw relevant conclusions from our results.

## 2 Mathematical Preliminaries

We give the following definitions that will be applied for proofs of the existence, uniqueness and positivity of the Caputo-Fabrizio (CF) model analysed in this work.

### Definition 1.

*[19] Assume ψ*(*t*) ∈ ℋ^1^(*ℓ*_1_, *ℓ*_2_), *for ℓ*_2_ *> ℓ*_1_, *p* ∈ [0, 1]. *Then the CF fractional operator is given as*

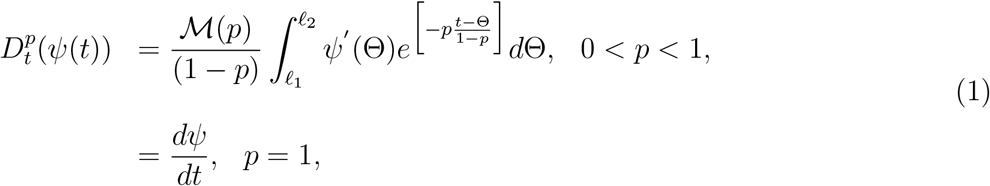

*where ℳ(p) satisfies the condition ℳ*(′) =ℳ(∞) = 1.

### Definition 2.

*The integral operator of fractional order corresponding to the CF fractional derivative defined in [13], which state*

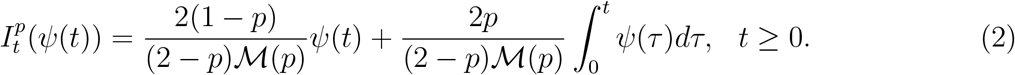

### Definition 3.

*The Laplace transform of* 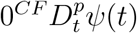 *is represented as follows*

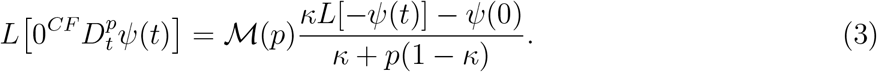

## 3 Fractional Dengue Fever Model Formulation

We modify the model by Olaniyi et al., [20] in which the total population is partitioned into the classes of susceptible humans *S*_*b*_(*t*), exposed humans *E*_*b*_(*t*), infectious humans *I*_*b*_(*t*), recovered humans *R*_*b*_(*t*), and the protected travellers *V*_*b*_(*t*). The total vector (mosquitoes) population is subdivided into susceptible mosquitoes *S*_*a*_(*t*) and infectious mosquitoes *I*_*a*_(*t*). Humans and mosquitoes are recruited into their respective susceptible classes at rates *π*_*b*_ and *π*_*a*_ respectively. Protected travellers are also recruited at a rate *ν*. Susceptible humans are bitten by mosquitoes at a rate *p* and they become infected and move into the exposed class. The probabilities that contact between a susceptible human and an infectious mosquito and between a susceptible mosquito and an infectious mosquito will result in infection are *β*_*b*_ and *β*_*a*_ respectively. Infected humans in the exposed class become infectious at a rate *δ*_*b*_ and can recover from Dengue fever at a rate *θ*. While in the recovered class, humans can either move into the class of protected travellers or lose their immunity and become susceptible to Dengue fever again. Individuals in all compartments die from natural causes at a rate *µ*_*b*_ while mosquitoes die naturally at a rate *µ*_*a*_.

The model diagram 1 and together with the model assumptions gives rise to the following system:

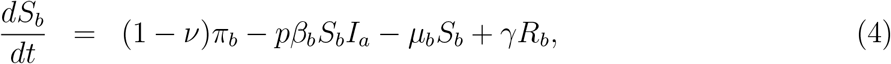

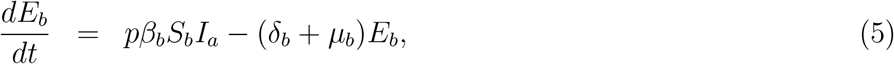

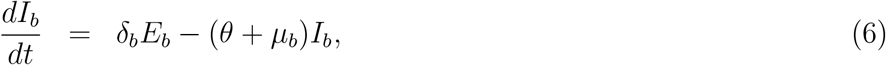

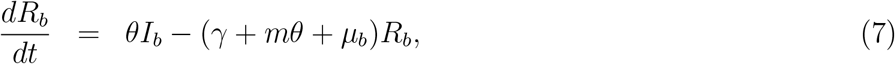

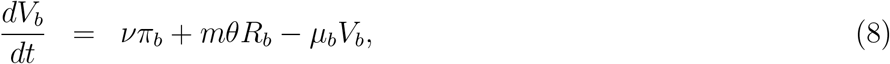

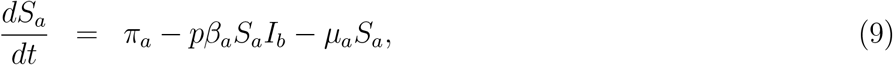

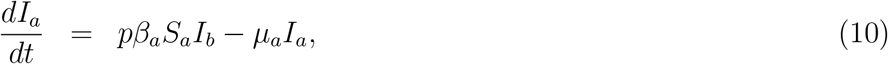

with initial conditions

*S*_*b*_(0) *>* 0, *E*_*b*_(0) ≥ 0, *I*_*b*_(0) ≥ 0, *R*_*b*_(0) ≥ 0, *V*_*b*_(0) ≥ 0, *S*_*a*_(0) *>* 0, *I*_*a*_(0) ≥ 0 for all *t* ≥ 0.

## 4 The Caputo-Fabrizio Model Properties and Analysis

In this section, we apply the Caputo-Fabrizio derivative to the Dengue fever model (4)-(10). The fractional order model of the Dengue fever incorporating protected travellers in the Caputo sense is given by:

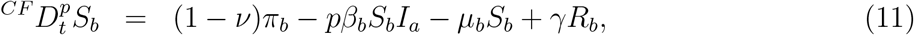

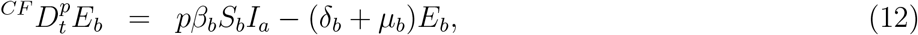

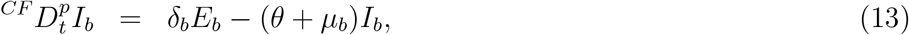

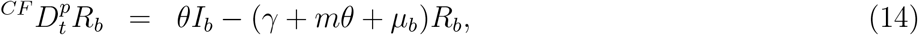

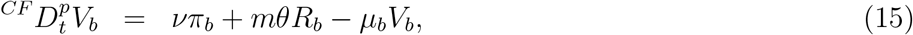

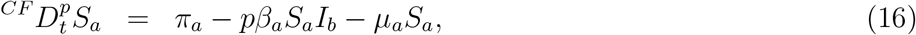

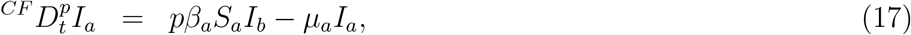

where *p* represents the fractional order, 0 *< p*, 1 and the initial conditions of the fractional order model are *S*_*b*_(0) *>* 0, *E*_*b*_(0) ≥ 0, *I*_*b*_(0) ≥ 0, *R*_*b*_(0) ≥ 0, *V*_*b*_(0) ≥ 0, *S*_*a*_(0) *>* 0, *I*_*a*_(0) ≥ 0, for all *t* ≥ 0.

### 4.1 Positivity of Solutions

#### Theorem 1.

*Given that S*_*b*_(0) *>* 0, *E*_*b*_(0) ≥ 0, *I*_*b*_(0) ≥ 0, *R*_*b*_(0) ≥ 0, *V*_*b*_(0) ≥ 0, *S*_*a*_(0) *>* 0, *I*_*a*_(0) ≥ 0, *for all t* ≥ 0, *we show that the set* 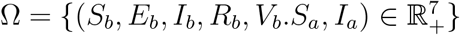 *attracts all positive solutions of the fractional order system (11)-(17)*.

We state the following lemma used in proving Theorem 1.

#### Lemma 1.

*[21] Suppose f* (*t*) ∈ *C*[*a, b*] *and* 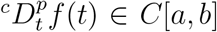 *for all* 0 *< p* ≤ 1, *then we have* 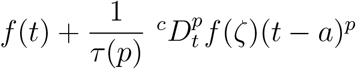, *where a* ≤ *ζ* ≤ *t, for all t* ∈ (*a, b*].

From Lemma 1, we have the following remark.

#### Remark 1.

*Assume that h*(*x*) ∈ *C*[*a, b*] *and* 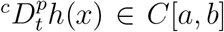 *for* 0 *< p* ≤ 1. *It follows from Lemma 1 that if* 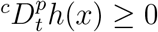, *for all x* ∈ (*a, b*), *then h*(*x*) *is non decreasing and if* 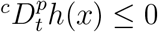 *for all x* ∈ (*a, b*), *then h*(*x*) *is non increasing*.

We now prove Theorem 1.

*Proof*. The existence and uniqueness of the solutions of the system (11)-(17) follow from Lemma 1 and Remark 1. We now prove that Ω is positively invariant by proving for each hyperplane bonding, the non-negative orthnant of the vector field points into Ω. From the system (11)-(17),

we have

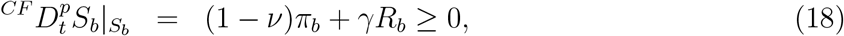

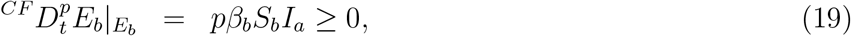

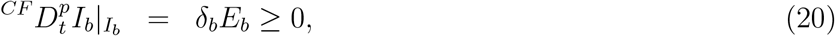

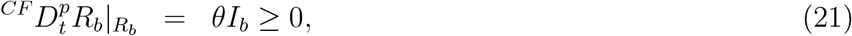

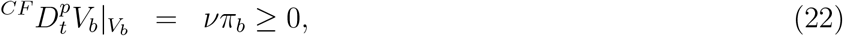

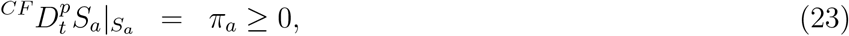

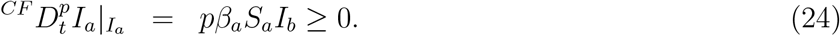

Therefore, Ω is positively invariant and attracts all positive solutions of the system (11)-(17) for *t* ≥ 0.

### 4.2 Existence and Uniqueness of Solutions

In this subsection, we investigate the existence and uniqueness of the solution of the fractional order system (11)-(17).

Applying the Losada and Nieto integral operator as in [13] to the system (4)-(10), we obtain

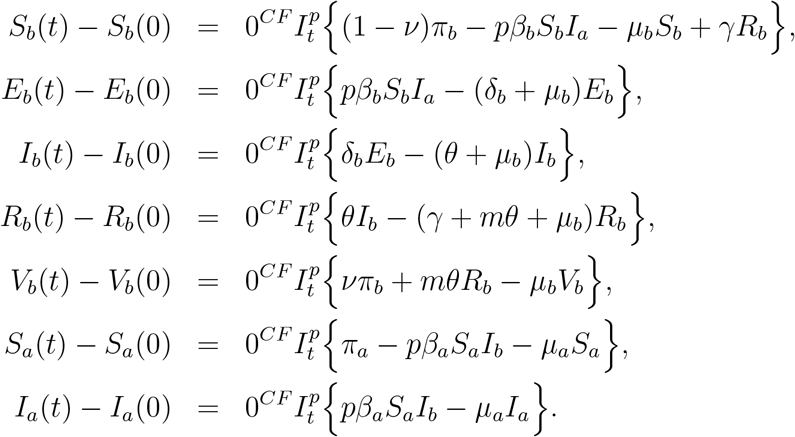

By the notation in [13], we have

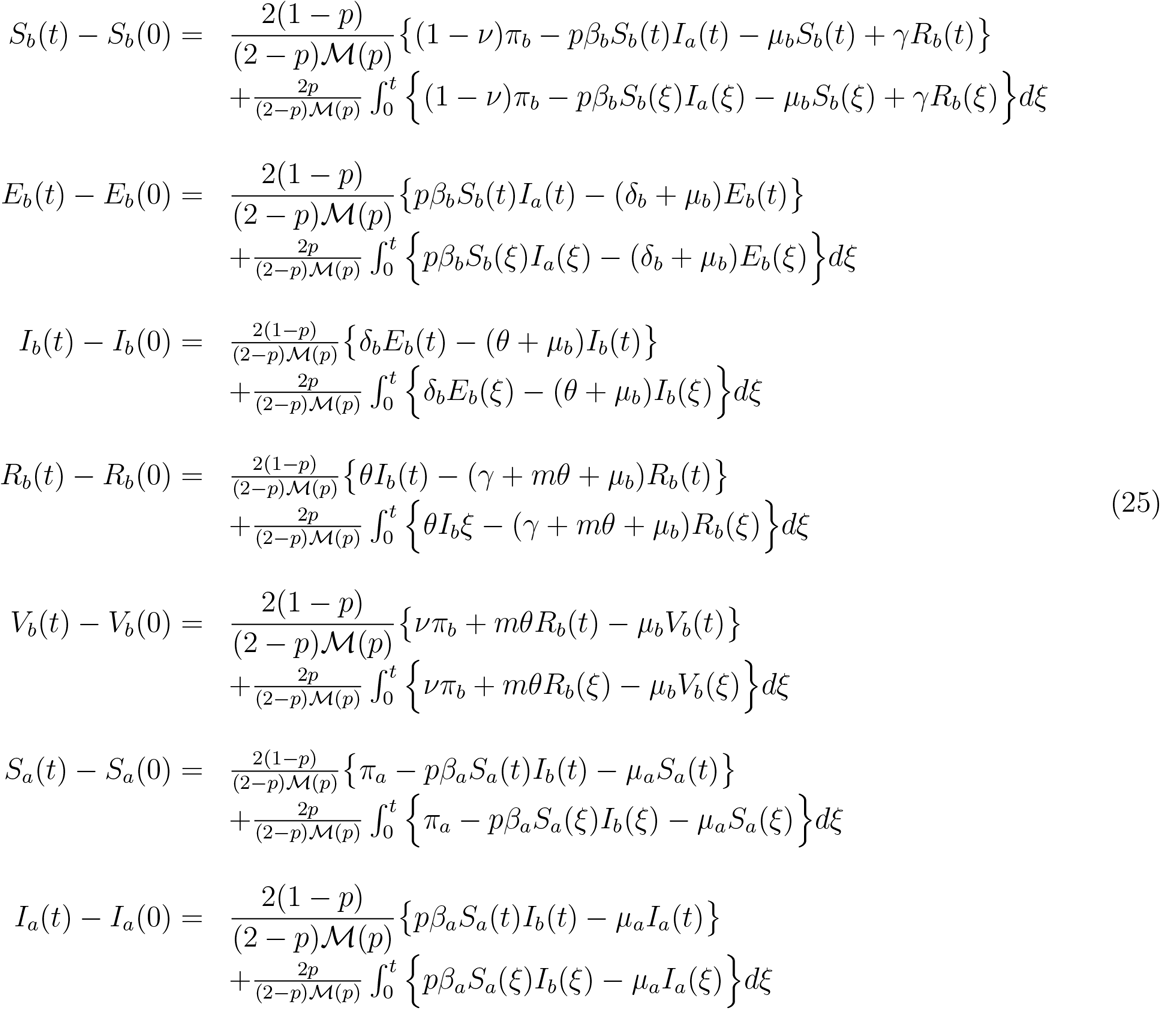

For the sake of simplicity, let

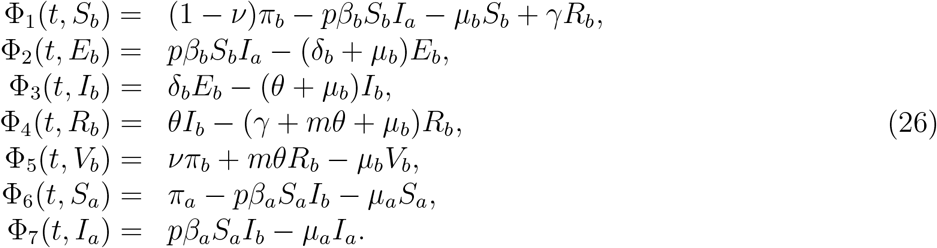

#### Theorem 2.

*The kernels* Φ_1_, Φ_2_, Φ_3_, Φ_4_, Φ_5_, Φ_6_ *and* Φ_7_ *satisfy the Lipschitz condition and contraction on if the following inequality holds if*

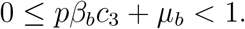

*Proof*. For a function *S*_*b*_ and *S*_*b*1_ 

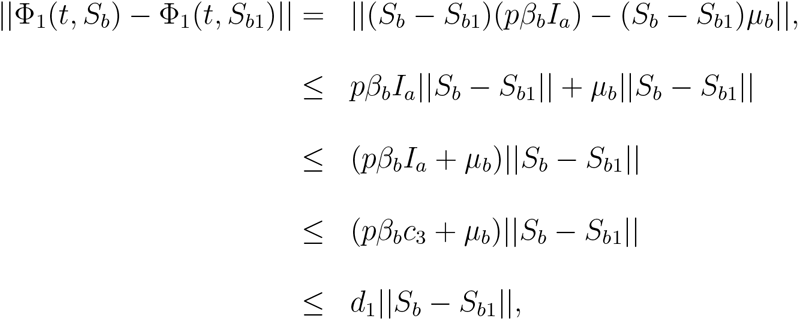

where *d*_1_ = *pβ*_*b*_*c*_3_ + *µ*_*b*_, ||*S*_*b*_(*t*)|| ≤ *c*_1_, ||*E*_*b*_(*t*)|| ≤ *c*_2_, ||*I*_*b*_(*t*)|| ≤ *c*_3_, ||*R*_*b*_(*t*)|| ≤ *c*_4_, ||*V*_*b*_(*t*)|| ≤ *c*_5_, ||*S*_*a*_(*t*)|| ≤ *c*_6_ and ||*I*_*a*_(*t*)|| ≤ *c*_7_ are bounded functions. Therefore,

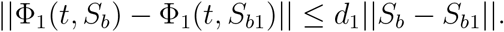

The Lipschitz condition is thus satisfied for Φ_1_ and if 0 ≤ *pβ*_*b*_*c*_3_ + *µ*_*b*_ *<* 1, then Φ is a contraction. Similarly we can prove that Φ_2_(*t, E*_*b*_), Φ_3_(*t, I*_*b*_), Φ_4_(*t, R*_*b*_), Φ_5_(*t, V*_*b*_), Φ_6_(*t, S*_*a*_) and Φ_7_(*t, I*_*a*_) satisfy the Lipschitz conditions.

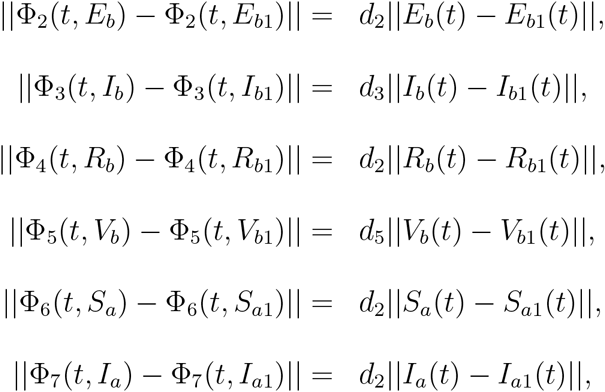

where *d*_2_ = (*S*_*b*_ + *µ*_*b*_)*c*_3_, *d*_3_ = *θ* + *µ*_*b*_, *d*_4_ = *γ* + *µ, d*_5_ = *µ*_*b*_, *d*_6_ = *pβ*_*a*_*c*_3_ + *µ*_*a*_ and *d*_7_ = *µ*_*a*_. Using the notation in equation (2) reduces (26) to the system

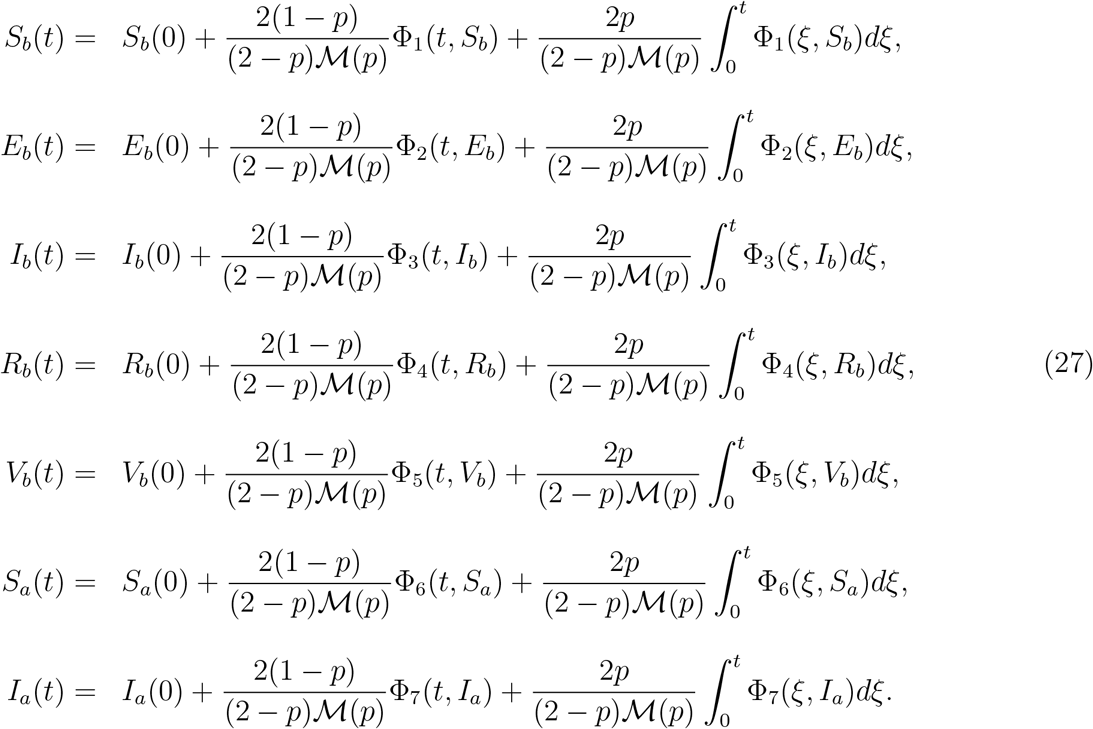

Consider the following recursive forms

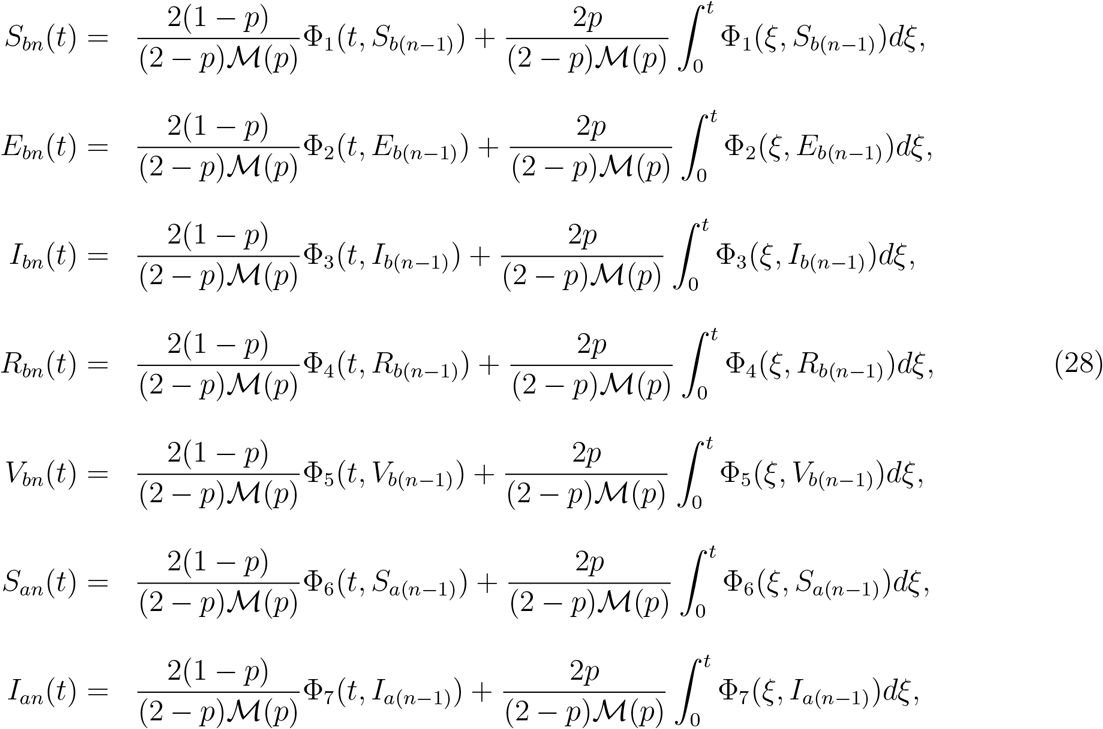

together with the initial conditions:

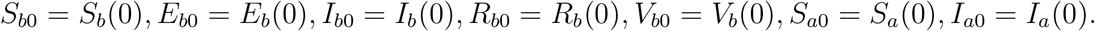

The difference between the succession terms is

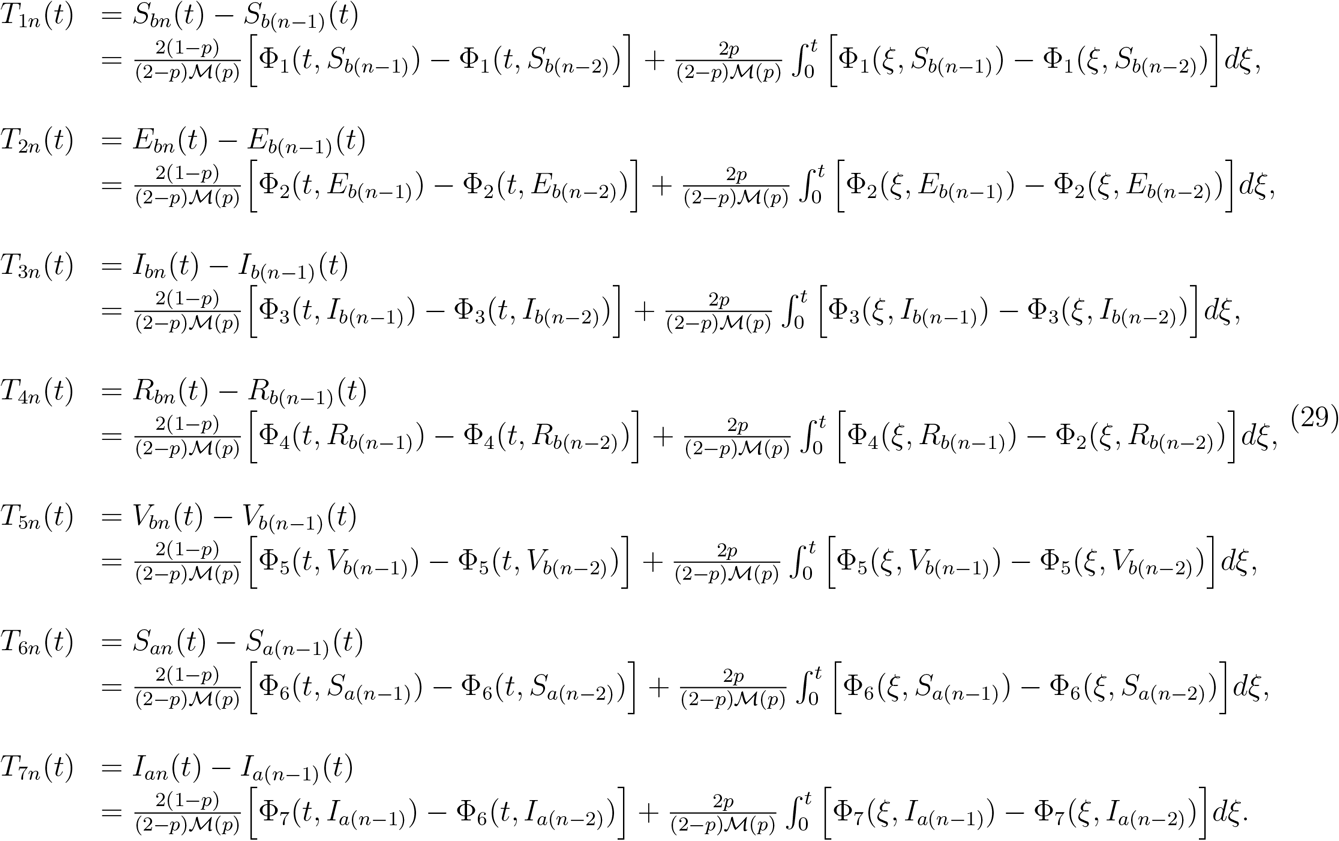

Observe that

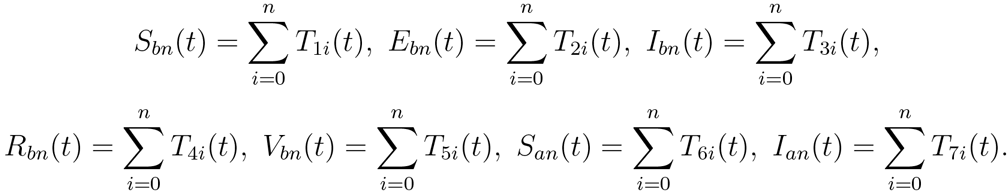

We thus have the following results

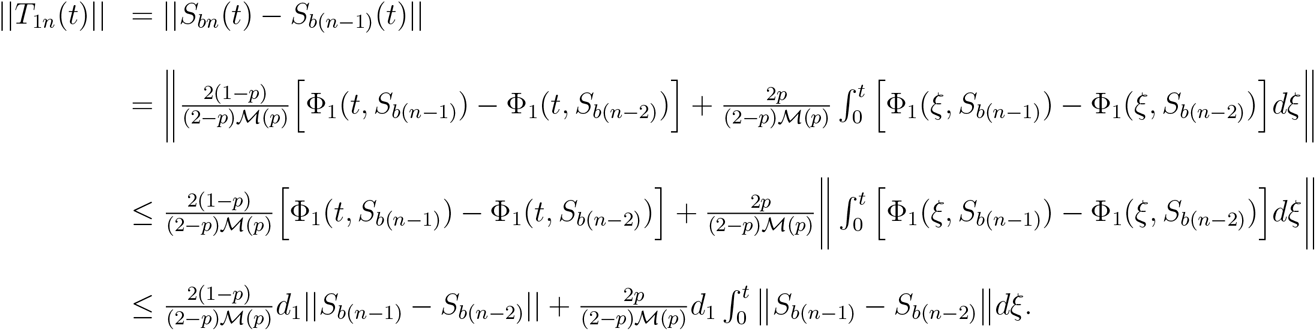

Thus,

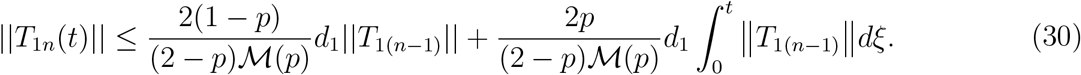

Similarly,

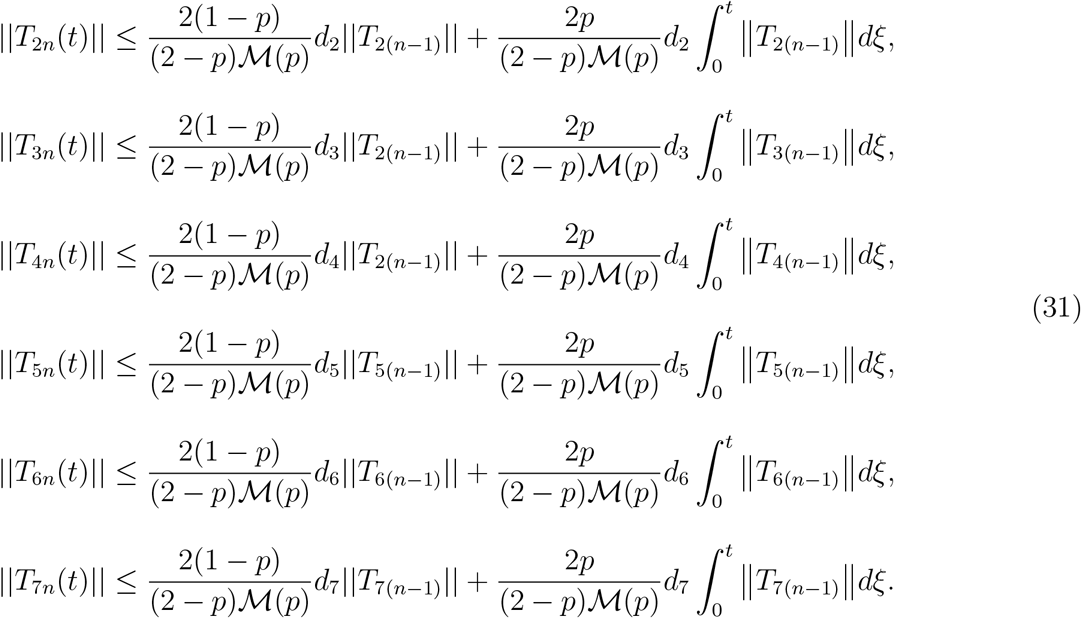

□

#### Theorem 3.

*The fractional order system (4)-(10) has a solution if there exists to such that* 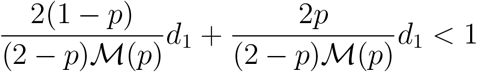.

*Proof. S*_*bn*_(*t*), *E*_*bn*_(*t*), *I*_*bn*_(*t*), *R*_*bn*_(*t*), *V*_*bn*_(*t*), *S*_*an*_(*t*) and *I*_*an*_(*t*) are all bounded functions. Using the results presented in equations (30) and (31) and the recursive algorithms, we have

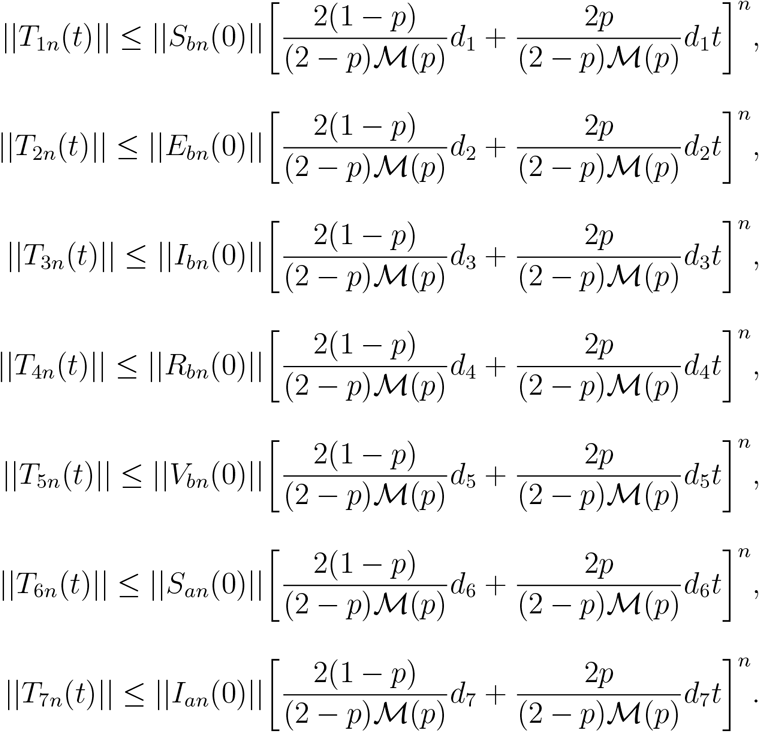

Hence, the solution of the model exists and is continuous. Next, we show that the solution is unique. Assume that the fractional model (4)-(10) has another solution 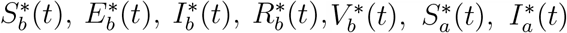, then,

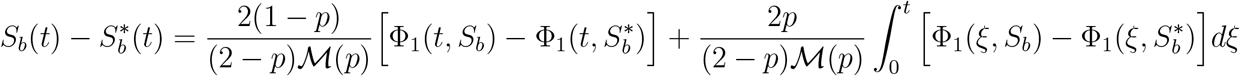

and

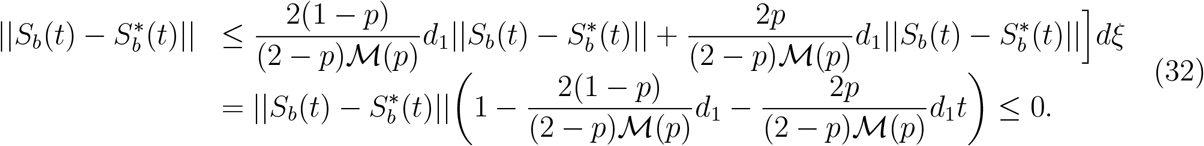

□

#### Theorem 4.

*The fractional order model (8)-(14) has a unique solution if*

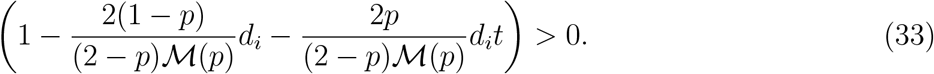

*Proof*. From equation (32), we have

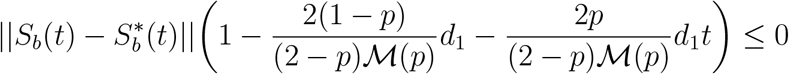

and from (33)

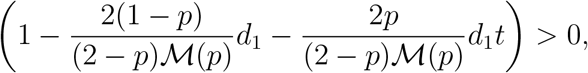

thus 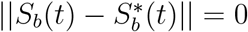, hence 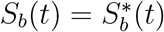. Similarly, we establish that 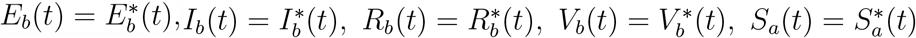 and 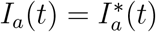.

### 4.3 Model Equilibria

#### 4.3.1 The Dengue Fever Free Equilibrium Point

In the absence of a Dengue fever infected individual or mosquito, that is 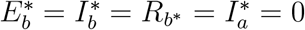. The system (11)-(17) yields

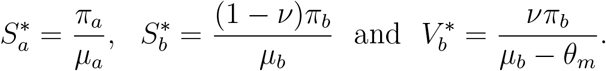

Therefore, the Dengue fever free equilibrium point denoted by *D*^0^ is given by:

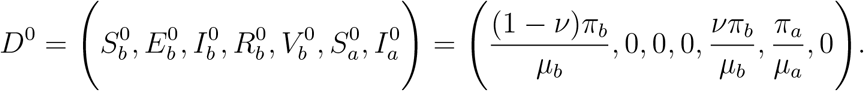

Before investigating the long term dynamics of the model, we first compute the basic reproduction number in the next subsection.

#### 4.3.2 The Basic Reproduction Number

We compute the basic reproduction number *R*_*D*_ using the next-generation matrix method [22] as follows. Let ℱ and 𝒱 be the new infections and transfer matrices of the model (11)-(17) respectively. (see [22] for the definitions of ℱ and 𝒱). The corresponding Jacobian matrices evaluated at the DFE point are:

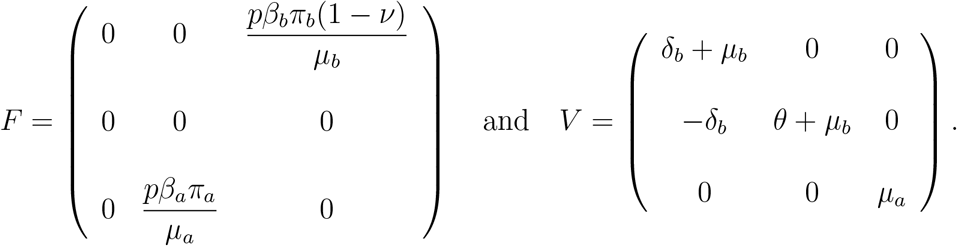

The reproduction number is the spectral radius of the next generation matrix, that is *FV* ^*−*1^, so that

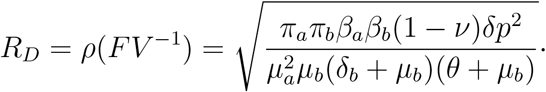

#### 4.3.3 The Dengue Fever Endemic Equilibrium Point

In this subsection, we present the endemic equilibrium solution of the fractional order dengue fever model (11)-(17) denoted by

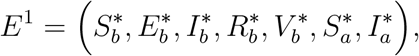

where

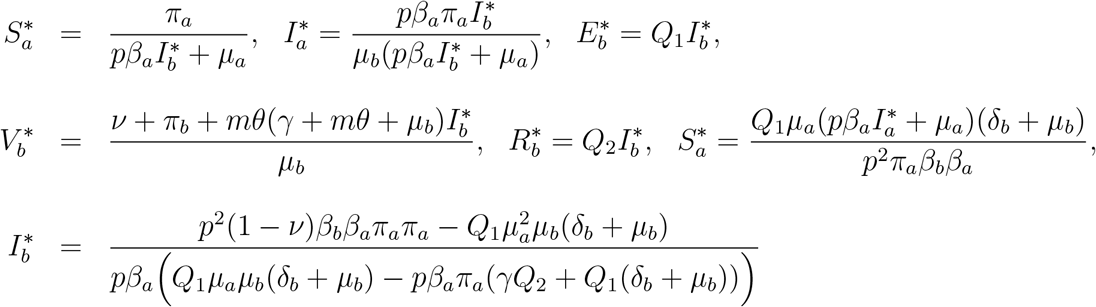

in which 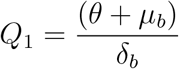 and 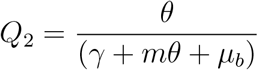.

### 4.4 Global Stability of the DFE

#### Theorem 5.

*The DFE is globally asymptotically stable in ω whenever R*_*D*_ *<* 1 *and unstable otherwise.*

*Proof*. We consider the positive definite *C*^1^ Layapunov function given by

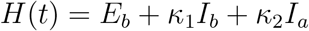

which consists of classes of individuals and mosquitoes who directly contribute to the disease transmission. *κ*_1_ and *κ*_2_ are non-negative constants to be determined. The CF derivative of *H*(*t*) is given by

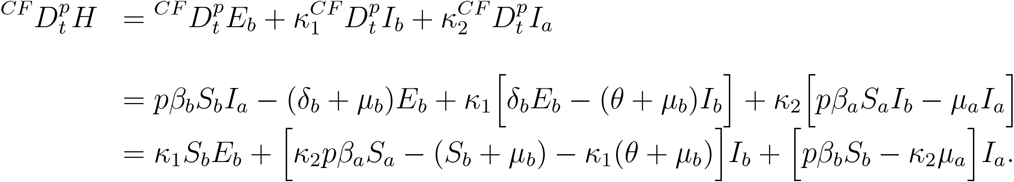

At DFE,

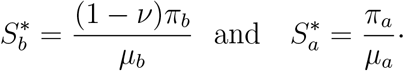

Therefore, the Lyapnouv function *H*(*t*) satisfies the inequality

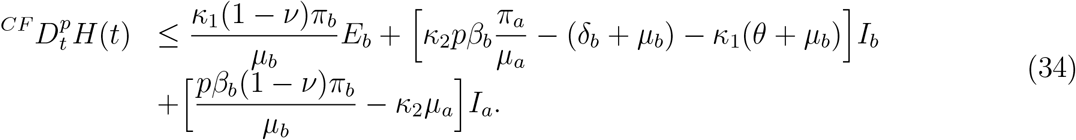

We equate the coefficients of *I*_*b*_ and *I*_*a*_ to zero and solve for *κ*_1_ and *κ*_2_ so that

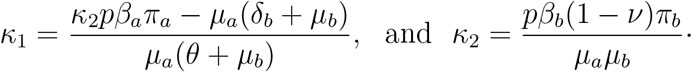

Substituting the constants into the inequality (34), we obtain 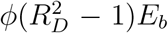 where *φ* = (*δ*_*b*_ + *µ*_*b*_)(1 − *ν*)*π*_*b*_. When *R*_*D*_ ≤ 1, 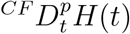 is negative semi definite with equality at *R*_*D*_ = 1, and *E*_*b*_ ∈ *E*^0^. Therefore, {*E*^0^} is the largest compact invariant set in Ω. Thus, by LaSalle’s invariant principle [23], the DFE is globally asymptotically stable in Ω if *R*_*D*_ ≤ 1 and unstable otherwise.

## 5 Numerical Simulations and Discussion

Here, we carry out numerical simulation of the dengue fever model based on the Caputo-Fabrizio operator. The Adams–Bashforth scheme as in [24] is employed in solving (11)-(17). The step size for the numerical simulations is 0.001 with time interval of [0,120]. The numerical simulation is based on the following parameter values.

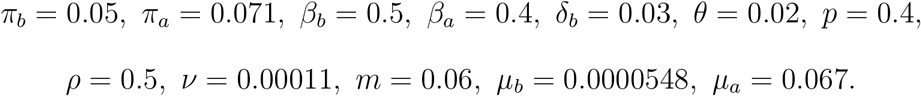

Figure 2(a) is the susceptible humans and as the fractional order is increased from 0.75 to 1 the number of susceptible humans’ decrease. As more humans move into the exposed class the susceptible population is expected to reduce in reality. In Figure 2(b) the number of exposed humans increase as the fractional order rise towards one. It can be inferred that as the number of exposed humans increase, the fractional-order also moves up. One major characteristic of exponential law is its crossover ability which ensures a better prediction. Thus the operator can stretch from one operator to another. Similarly, in Figure 2(c) as the fractional order increases, the number of humans infected with dengue fever go up. Naturally, this is expected in a typical epidemiological model. Figure 2(d) is the recovered humans but as the fractional order increases, the number of recovered humans also decrease. Therefore, one can say that, to reverse the situation in the community, efforts must be made to reduce the fractional-order so that more humans can recover. Thus, there should be more efforts towards treatment and boosting the immunity of the infected humans. In Figure 3(a) the number of protected travellers increase as the fractional order also increases. This means that effort must be made to push up the fractional-order to have as many protected travellers as possible. Figure 3(b) depicts the number of susceptible mosquitoes in the communities and as the fractional order increases the number of susceptible individuals decrease. This is naturally expected in an epidemiological model because they move to the infected compartment as more get infected. Figure 3(c) is the infectious mosquitoes and as the fractional order increases the number of infectious mosquitoes also go up. For the infectious mosquitoes to be minimized effort must be made to reduce the fractional order.

**Figure 1:**
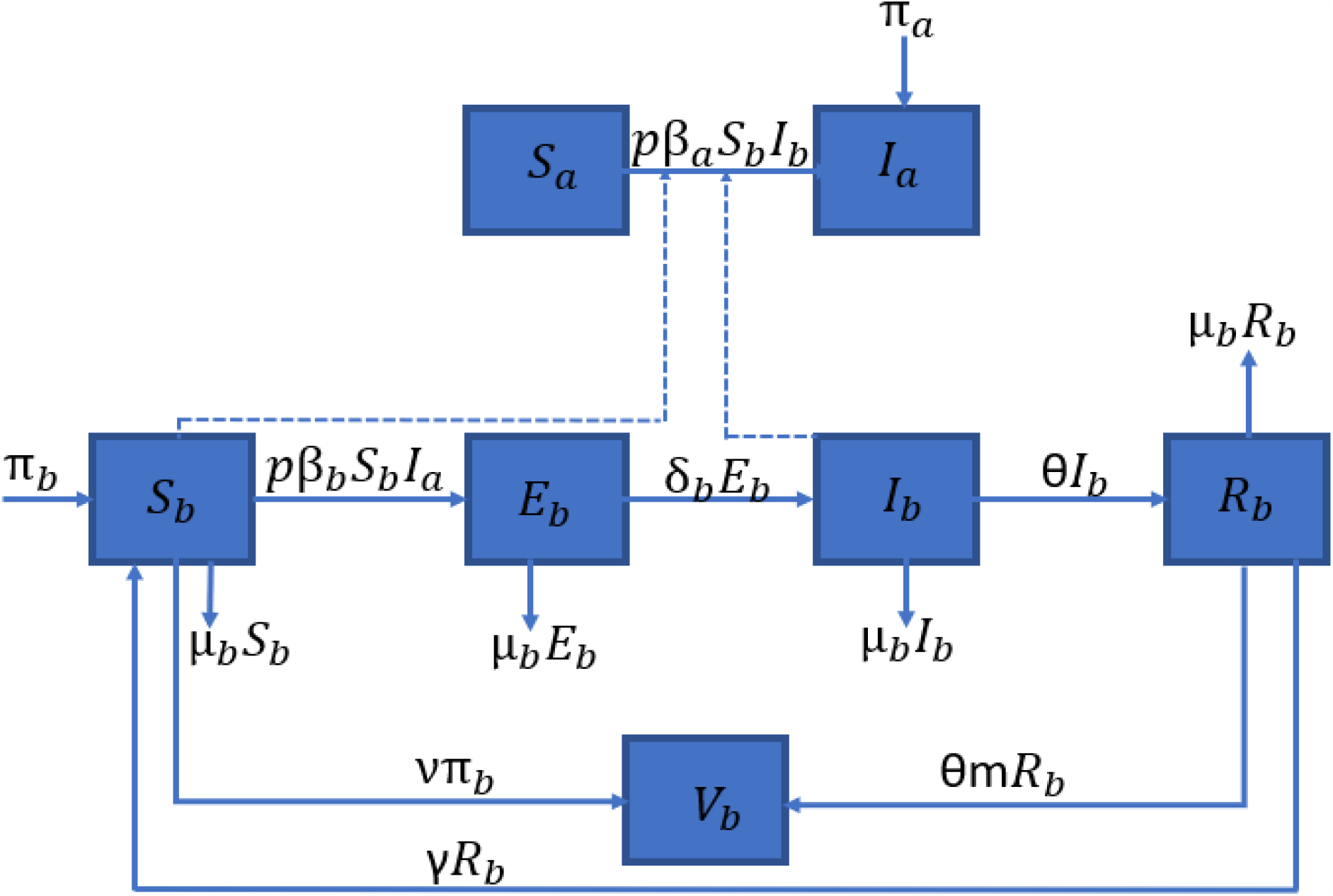
Dengue fever model flow diagram describing the disease dynamics in human population (in the context of protected travellers context) by a vector.

**Figure 2:**
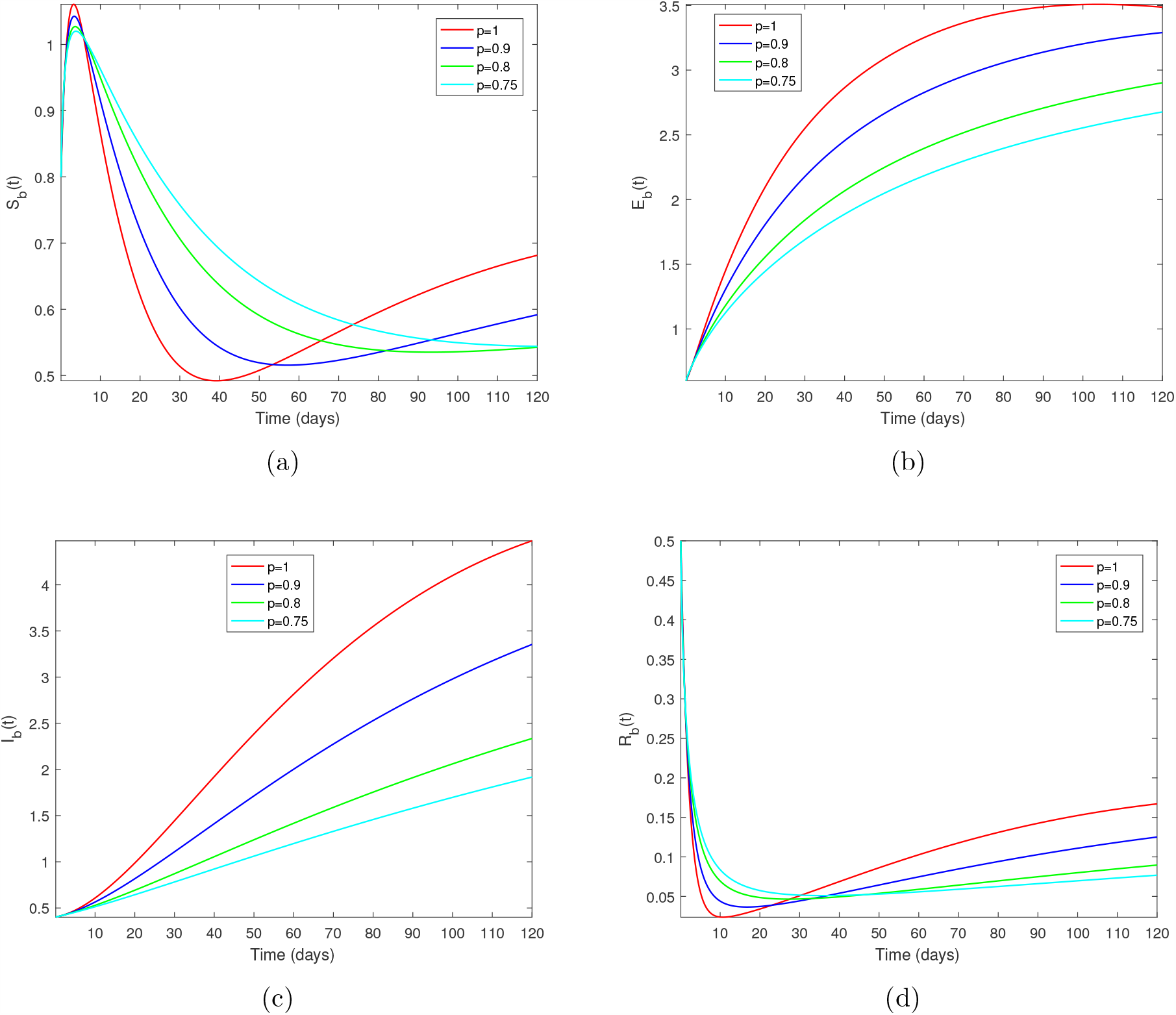
Simulations for Dengue fever model (11)-(17) via Exponential-law at p = 1, 0.9, 0.8, 0.75.

**Figure 3:**
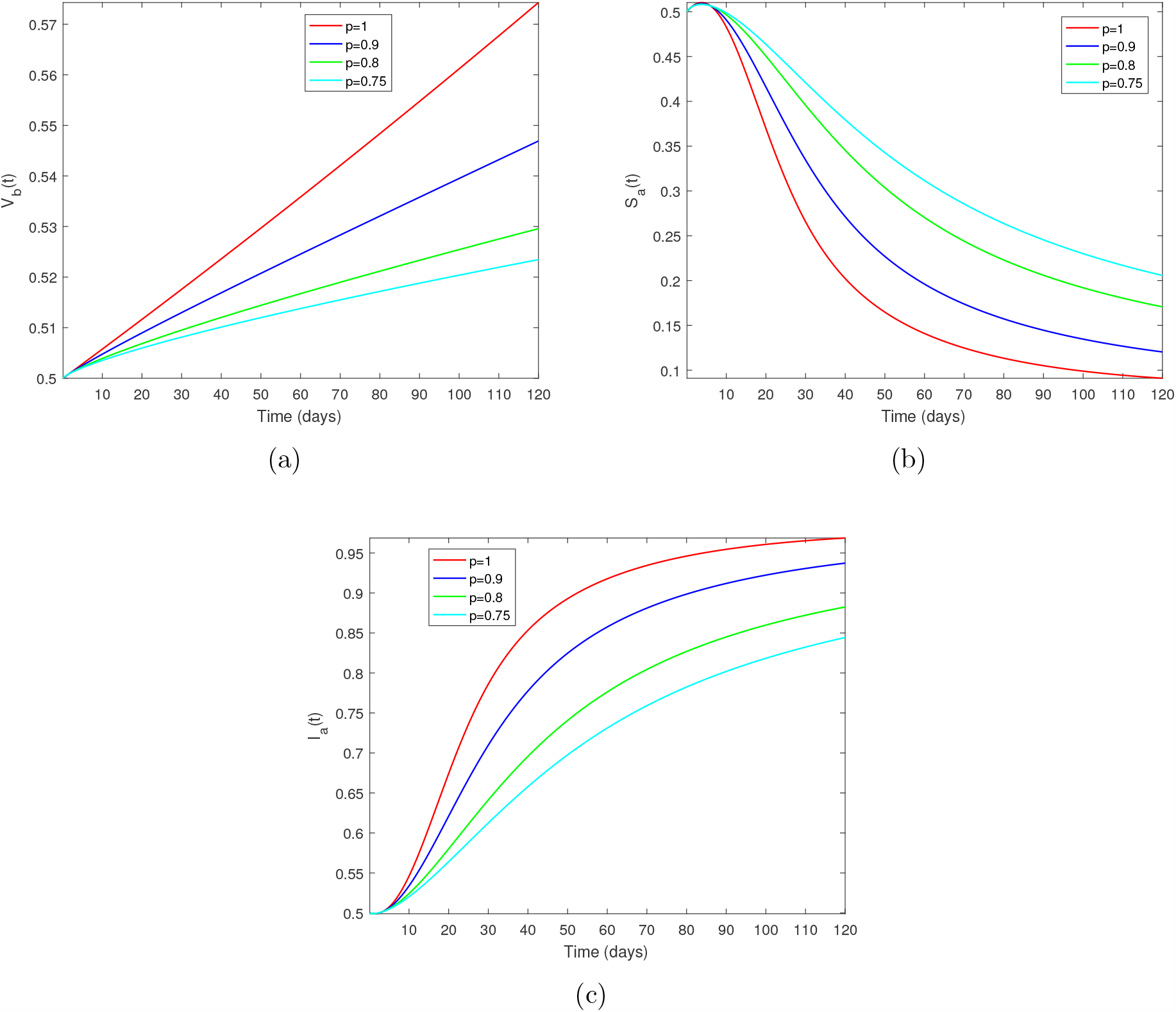
Simulations for Dengue fever model (11)-(17) via Exponential-law at p = 1, 0.9, 0.8, 0.75.

## 6 Conclusion

In this paper, the dengue fever model with travellers’ protection was formulated and analysed. Fractional derivatives based on the exponential decay law approach was utilized for the analysis. The basic properties of the model have been investigated as well as the steady states. The stability analysis of the dengue fever model was examined and found to be globally stable at the disease free state. Fixed point theory was employed to determine the existence and uniqueness of solutions of the dengue fever model. The numerical simulation result shows that fractional-order affects the dynamics of the dengue fever model. In conclusion, complex biological models can be explored using the exponential law.

## Data Availability

None

## Acknowledgements

Authors will like to say thanks to their respective Universities for the production of this manuscript.

